# Effect of health-system continuous quality improvement on long-term retention in HIV care among pregnant and breastfeeding women in the Democratic Republic of Congo: A cluster-randomized trial

**DOI:** 10.64898/2026.09.24.26363877

**Authors:** Marcel Yotebieng, Noro Lantoniaina Rosa Ravelomanana, Bienvenu Kawende, Martine Tabala, Fathy Malongo, Fidéle Lumande, Natalia Zotova, Pelagie Babakazo

## Abstract

**Background:** Despite relatively high coverage of interventions to prevent vertical transmission, HIV incidence among children born to women living with HIV (WLH) remains above elimination targets, partly because of substantial disengagement from antiretroviral therapy (ART) during the postpartum period. We evaluated the effectiveness of a data-driven continuous quality improvement (CQI) intervention in improving retention in care, virological suppression, and prevention of vertical transmission among pregnant and breastfeeding WLH receiving ART in maternal and child health (MCH) clinics.

**Methods and findings:** This pragmatic cluster-randomized trial was conducted in 105 MCH clinics— three high-volume clinics in each of the 35 health zones (HZs) of Kinshasa Province, Democratic Republic of the Congo. During the pre-randomization phase (November 2016–December 2017), routine data collection using prevention of mother-to-child transmission registers was strengthened in all participating clinics. In January 2018, HZs were randomized 1:1 to implement CQI or continue the standard of care. In intervention HZs, quality improvement (QI) teams were established at the HZ and clinic levels. Indicators selected collaboratively with QI teams were calculated from routine program data and shared quarterly. Through July 2019, QI teams met quarterly to identify service-delivery challenges and develop strategies to improve outcomes, with an emphasis on retention. Thereafter, responsibility for producing indicators was transferred to QI teams, and meetings continued virtually through WhatsApp.

Pregnant and breastfeeding WLH and their infants receiving care at participating clinics between November 2016 and July 2019 were enrolled and followed for at least 24 months. Primary outcomes were loss to follow-up (LTFU), viral suppression (<1,000 copies/mL), and a positive HIV test among live-born infants. Overall, 2,376 women were enrolled, including 1,217 (51.2%) before randomization. The analysis of CQI effectiveness included 1,900 participants who remained in care at randomization or enrolled thereafter: 1,103 (58.1%) in control HZs and 797 (41.9%) in intervention HZs.

At 6, 12, and 24 months, LTFU was lower in the intervention group than in the control group: 18.6% versus 21.6% (adjusted risk difference [aRD], −5.2 percentage points; 95% CI, −9.6 to −0.8), 29.2% versus 31.0% (aRD, −3.9; 95% CI, −10.2 to 2.4), and 50.1% versus 55.5% (aRD, −7.1; 95% CI, −15.2 to 1.0), respectively. Estimated effects were larger in PEPFAR-supported and urban clinics, although interaction tests were not statistically significant. Viral-load results were available for 38.2%, 50.4%, and 73.5% of eligible participants at 6, 12, and 24 months, respectively. Neither viral-load-result availability nor viral suppression differed between groups. Infant HIV-test results were available for 8.1%, 32.7%, 34.2%, and 28.5% of eligible infants at 6 weeks and 6, 12, and 24 months, respectively.

**Conclusions:** CQI implementation produced a modest improvement in retention, with the clearest evidence at 6 months and favorable but imprecise estimates through 24 months. Limited availability of maternal viral-load and infant HIV-test results precluded definitive conclusions about virological suppression and vertical transmission. Local quality-improvement interventions may strengthen processes within frontline teams’ control but cannot alone overcome failures across an interconnected care system. Future studies should align interventions’ operational reach with all health-system levels responsible for the intended outcomes.

**Trial registration:** http://ClinicalTrials.gov <u>NCT03048669</u>.

**Why was this study done?:**

- Loss from HIV care during pregnancy and breastfeeding remains an important barrier to preventing HIV transmission from women to their children.
- Most interventions have focused on supporting individual women, while relatively few have tested approaches that help clinics and local health authorities identify and correct weaknesses in healthcare delivery.
- Randomized evidence about whether continuous quality improvement can improve long-term retention in maternal HIV care is limited, particularly in resource-constrained settings.

**What did the researchers do and find?:**

- We conducted a cluster-randomized trial involving 105 maternal and child health clinics across all 35 health zones in Kinshasa, Democratic Republic of Congo; the intervention-period analysis included 1,900 women living with HIV.
- In intervention health zones, clinic and health-zone teams regularly reviewed performance data, identified barriers to care, and implemented locally selected solutions; comparison health zones continued standard care.
- Loss to follow-up was approximately 5 percentage points lower at 6 months, 4 percentage points lower at 12 months, and 7 percentage points lower at 24 months in the intervention group after adjustment, although only the 6-month estimate clearly excluded no effect.
- Viral suppression and infant HIV outcomes did not differ between groups, but these analyses were limited by the low availability of laboratory results.

**What do these findings mean?:**

- Data-driven continuous quality improvement may produce modest improvements in retention when frontline clinic and health-zone teams can directly act on the barriers responsible for poor outcomes.
- Local quality-improvement activities may have less effect on outcomes that depend on services outside their operational control, such as testing and reporting performed through a centralized laboratory system.
- The longer-term estimates were imprecise, implementation varied among clinics, and many laboratory results were unavailable; the findings should therefore be interpreted cautiously and confirmed in other health systems.

## INTRODUCTION

Globally, about 1 million women living with HIV (WLH) became pregnant in 2024[1]. In absence of any intervention, pregnant or post-partum WLH have around eight times higher mortality than their HIV-uninfected counterparts[2]; 20−45% of them will transmit the virus to their infants through mother-to-child transmission (MTCT)[3]; and one third of those infants infected perinatal will not survive their first birthday [4]. Early initiation of triple-antiretroviral therapy (ART) that results in sustained viral suppression delay the progression of disease in both adults [5] and infants [6], and enables them to live longer [7], and reduce the risk of transmission [8].

Since 2010, when the World Health Organization (WHO) first recommended ART for pregnant women to prevent vertical transmission (VT) [9], new HIV infections among children have declined by 62%, from 310 000 to 120 000 in 2024 [10]. In this era of universal ART, ambitious targets for ending the AIDS epidemic has been set including a reduction in the number of children 0-14 years of age newly infected with HIV from 150,000 in 2022 to 20,000 in 2025 and 15,000 in 2030 [11]. However, evidence from a recent systematic review and meta-analysis to update estimates of VT probabilities [12], showed that, despite the relatively high coverage of ART among pregnant WLH, the VT probability remains high, primarily because of high ART drop-off [12], highlighting the persistent need for strategies to address LTFU in the postpartum period.

In fact, since the introduction of interventions to prevent VT that require longitudinal follow-up of women throughout pregnancy and beyond, LTFU has been a major challenge to its effectiveness [13,14]. Consequently, numerous strategies have been designed and implemented to limit this drop-off including text messaging, peer’s support, community health workers, conditional cash transfer among others[15,16]. But they all virtually focus on WLH (patients) ignoring two other critical components of the health service triad: the health care organization and clinicians (team of physicians, nurses, medical assistants, and office staff).

Data-driven continuous quality improvement (CQI) is a systematic approach to enhancing organizational excellence that leverages data to identify areas for improvement, implement targeted interventions, and track progress. Since 2010, WHO has called for the development and promotion of quality improvement methods to strengthen regional and district-level health systems to improve the quality and reliability of HIV prevention, care and treatment services for women and children [17]. However, despite multiple observational studies suggesting that quality improvements are associated with improved outcomes [18–21], few randomized controlled trials have evaluated its effect on LTFU.

To the best of our knowledge only two trials have evaluated the effectiveness of CQI interventions on quality of HIV care. In a cluster randomized trial involving 26 facilities and 511 WLH in Nigeria, the authors reported a small non-statistically significant increase in retention in the CQI group relative to control (44% vs. 41%) at 6 months postpartum [22]. But in addition to the small sample size, the follow-up of 6 months is relatively short and in most settings of high VT burden, breastfeeding continues well beyond 6 months. In a stepped-wedge cluster-randomized controlled implementation trial to evaluate the effectiveness of continuous quality improvement (CQI) on viral load (VL) monitoring and repeat HIV testing in peri-urban South Africa, CQI implementation was found to significantly increase VL monitoring (relative risk [RR] 1.38, 95% CI 1.21 to 1.57, p < 0.001) but did not improve repeat HIV testing[23]. Thus, whether CQI can be used to address the issue of long-term retention in prevention of vertical transmission programs has not been evaluated.

The aim of this study was to evaluate the effectiveness of data-driven CQI implementation on LTFU and potential impact on virological suppression, and vertical transmission rate in pregnant and breastfeeding WLH receiving ART in MCH clinics in Kinshasa, the Democratic Republic of Congo (DRC).

## Methods

### Study settings

The study was implemented in Kinshasa, the capital city province of the Democratic Republic of Congo (DRC). In Kinshasa, as part of the prevention of VT program, all women presenting for antenatal care (ANC) with unknown HIV status are tested for HIV and, since 2015, all pregnant women with HIV are immediately initiated on ART. A mother-infant register – modeled after the WHO’s *Three Interlinked Patient Monitoring Systems for HIV Care/ART, MCH/PMTCT (including malaria prevention during pregnancy), and TB/HIV: Standardized Minimum Data Set and Illustrative Tools* 24] – is used to track important HIV care events among WLH and their infants from ART registration until the end of postnatal visits 18-24 months after delivery. At the end of the 18-24 months, WLH who are not pregnant again and their infants (if HIV-infected) are referred to the HIV clinics for the continuation of their HIV care. At least every quarter, the PMTCT team at the clinic uses data from the register to calculate key PMTCT indicators and report them to the Health Zone bureau, usually during a supervising visit from the bureau to the clinic. Sample for HIV testing is collected from children born to WLH at 6 weeks for early HIV diagnosis and at subsequent visits to at least 3 months after cessation of all breastfeeding. The samples are sent to the national laboratory for processing and testing. Viral load testing is recommended during pregnancy and at least once a year. Similar to infant HIV testing, blood samples for Viral load were processed and tested centrally at the national laboratory, run by the National AIDS program.

### Study design

The study was designed to be implemented in two phases. During the first phase, we obtained from the National AIDS Control Program a list of 304 maternal and child health (MCH) clinics across the 35 Health Zone in the Kinshasa province implementing PMTCT interventions and selected 128 with the largest number pregnant served in 2015. Between July 2016-December 2017, study staff visited and surveyed each of 128 clinics and the confirmed top three in terms of number of pregnant WLH served in 2015 and 2016 were selected for the study. Starting January 2017, every month, study personnel visited each clinic to extract the data from the register and worked with the clinic staff to ensure completeness of the data. Phase II started with randomization in January 2018.

### Randomization

All 35 Health Zones in Kinshasa were randomized 1:1 to CQI or standard of care using the list of Health Zones. Although the interventions were implemented mainly at the clinic level, the health zone was chosen as the unit of randomization because the first level of supervision for MCH clinics is the Health Zone Bureau, to avoid potential contamination of the control clinics. By the nature of the intervention, it was not possible to mask the intervention to clinics and study staff. However, participants (WLH) were not informed about the randomization nor the intervention.

### Interventions

In Health Zones randomized to intervention, a quality improvement team was set up at the level of the Health Zone that includes the PMTCT supervisor from the Health Zone Bureau and at least the head of PMTCT from each of the participating MCH clinics. In each clinic, a clinic level QI team was also established and included at least one staff member each from ANC, delivery/maternity, and well-child services. A study staff member served as rapporteur of each QI team. Immediately following randomization, QI teams were brought together for a half-day meeting during which they reviewed key indicators that were calculated using data from registries and identified key bottlenecks in the care delivery system affecting those indicators, developed an action plan to address those bottlenecks and defined indicators to monitor progress to be included in the next quarterly report. The choice of indicators and action plan was done collaboratively with the input from everyone from the clinic to the health zone bureau. Clinic QI teams were responsible for the implementation of the action plan at the level of their respective clinics. During their routine visit to the clinic, a member of the health zone QI team provided supervision and support for the implementation of the action plan including facilitation of access to supplies that may be in limited quantity at the clinic. Every three months, using data from the monitoring system, the study team generated a color-coded dashboard tracking progress of the selected indicators using the ‘traffic light’ approach. An indicator was scored green when the target had been achieved, yellow/amber when there was progress, but the target was not reached, and red when there was no progress or downward movement. The report was generated for each participating clinic in the intervention group and shared with the QI teams. The cycle was repeated through July 2019, after which period, quarterly meetings were switched from in-person to virtual using WhatsApp, and the task of producing indicators was transferred to the health zone and the clinics. To limit possible contamination, all study staff assigned to randomized Health Zones/clinics were restricted to those sites. To assess how well the continuous quality interventions were integrated in each facility, in January and February 2019, each of the 51 facilities was surveyed to assess the effectiveness of CQI meetings, including its documentation.

### Standard of care

In the standard of care group, activities continued as usual. No report on indicators was produced for the duration of the study by the study team. Staff from clinics and health zone bureaus in the standard of care group were not associated with the quarterly review of the indicators. The study did not interfere with any other HIV service provision activity in the standard of care group.

### Study population

All pregnant and breastfeeding WLH receiving HIV care at any selected clinics at the time of randomization January 2018 or newly enrolled in care at the site between randomization and July 2019 and consented to participate were eligible. Participants were excluded if they had given birth more than 12 months prior. All eligible participants were approached during a routine visit to the clinic and those who consented to be part of the study were enrolled and followed up till they dropped out, were transferred out, died, or the study closure in July 2021, whichever one occurred first.

### Outcomes

Two primary outcomes were considered: loss-to-follow-up (the proportion of participants for whom the whereabouts was unknown at the study closure) at 6, 12, and 24 months from randomization or enrollment (for those enrolled after randomization) and virological suppression (proportion of participants with viral load <1000 copies/ml at 0, 6, 12, and 24 months after randomization/enrollment). Participants who interrupted care for any reason and reengaged prior to study closure were classified as in-care. Data on two secondary outcomes are also presented here: 1) Timely infant HIV diagnosis (Proportion of HIV-exposed infants with an appropriate HIV test result at 6 weeks, 12 and 24 months postpartum) and 2) MTCT rates (proportion of infants born to participating women who tested positive for HIV at 6 weeks, 12 and 24 months postpartum).

### Statistical analysis

Participant characteristics were summarized by randomized group using medians and interquartile ranges for continuous variables and frequencies and percentages for categorical variables. Because this was a cluster-randomized trial, baseline characteristics were described to assess the magnitude and direction of between-group imbalances rather than to determine eligibility for covariate adjustment on the basis of statistical significance.

The primary analysis followed the intention-to-treat principle: participants were analyzed according to the intervention assigned to their health zone, regardless of subsequent transfer of care or the extent to which their clinic implemented the intervention. We estimated the proportion of participants lost to follow-up at 6, 12, and 24 months after January 1, 2018, for participants already receiving care at randomization, or after enrollment for participants enrolled subsequently. Binomial regression models with an identity link were used to estimate absolute risk differences and corresponding 95% confidence intervals comparing the CQI and standard-care groups. Generalized estimating equations with facilities nested within health zone as the clustering unit, an exchangeable working correlation structure were used to account for the clustered design. Adjusted models included marital status, educational attainment, employment status, primigravida status, facility management type, urban versus peri-urban location, and receipt of technical support from a PEPFAR implementing partner. These variables were included because of their prespecified prognostic importance and observed between-group imbalance.

The effects of CQI on viral suppression were estimated using the same modeling approach. Viral-suppression analyses were restricted to participants with an available viral-load result within the prespecified window at each evaluation time point. No imputation was performed for missing viral-load or infant HIV-test results; consequently, estimates for these outcomes represent complete-case analyses conditional on result availability. The availability of viral-load and infant HIV-test results was also compared between randomized groups. Infant HIV positivity was summarized among infants with an available valid result. Because of the substantial amount of missing laboratory data, analyses of viral suppression and infant HIV outcomes were interpreted as secondary and exploratory.

We conducted exploratory subgroup analyses according to facility location (urban versus peri-urban) and receipt of technical support from a PEPFAR implementing partner. For each potential effect modifier, we fitted models containing intervention group, subgroup, and intervention-by-subgroup interaction terms. Subgroup-specific risk differences and 95% confidence intervals were estimated from these models. Statistical evidence of effect modification was assessed using the interaction term rather than the statistical significance of estimates within individual subgroups. These analyses were not adjusted for multiple comparisons and were interpreted cautiously.

In a secondary analysis, we compared the cumulative incidence of loss to follow-up between randomized groups, treating death and transfer out as competing events. The preintervention analysis included participants enrolled before randomization; follow-up was censored at death, transfer out, or December 31, 2017, whichever occurred first. The intervention-period analysis included participants who remained in care on January 1, 2018, and participants enrolled subsequently. Follow-up began on January 1, 2018, or the date of enrollment, whichever occurred later. Cumulative incidence functions were compared using Gray’s test.

To quantify within-facility clustering of loss to follow-up, we fitted separate logistic generalized linear mixed-effects models at 6, 12, and 24 months, with a random intercept for facility nested within health zone. Facility-level intraclass correlation coefficients were calculated on the latent logistic scale as 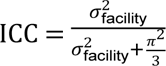, where 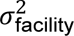 is the estimated facility-level random-intercept variance and *π*^2^/3 is the assumed individual-level residual variance. Confidence intervals for the ICCs were obtained by transforming the confidence limits for the corresponding variance components.

All statistical tests were two-sided, and p values below 0.05 were considered statistically significant. Effect estimates are presented with 95% confidence intervals. Analyses were conducted using SAS version 9.3 (SAS Institute, Cary, North Carolina, USA).

Sample-size requirements for the parent study were based on the planned multivariable analyses. We used a criterion of seven outcome events per candidate predictor parameter. The initial models were expected to contain 15 parameters: one contextual-level factor, two clinic-level factors, seven patient-level factors, and five additional terms for potential nonlinear trends or predictor-by-time interactions. Thus, at least 105 outcome events were required. Assuming that 20% of participants would be lost-to-follow-up during the first six weeks and a further 5% during each subsequent year, approximately 378 participants would provide sufficient events for the loss-to-follow-up analysis. The larger enrollment target was determined by the analysis of mother-to-child HIV transmission, for which the cumulative incidence was expected to be 5% by 24 months. Observing 105 transmission events would require 2,100 mother–infant pairs with complete follow-up. After allowing for the anticipated retention proportions of 80% through six weeks and 95% during each of the following two years, the enrollment target was 2,909 mother–infant pairs (2,100/[0.80 × 0.95 × 0.95]). Assuming that each eligible clinic would identify approximately 10 pregnant women living with HIV annually, the study planned to recruit participants from approximately 105 maternal and child health clinics—three clinics in each of 35 health zones—during the first three years.

### Ethical consideration

The study was approved by The Ohio State University institutional review board (Study ID: 2015H0440) and the Kinshasa School of Public Health Ethical committee. All participants provided signed informed consent. At the time the study initiation, pregnant people <18 years of age were considered emancipated minor in DRC. Parental permission was obtained before enrollment of HIV-exposed infants. The protocol was registered to ClinicalTrials.gov: NCT03048669.

### Role of the funding source

The funder of the study had no role in the study design, data collection, data analysis, data interpretation, or writing of the report. The corresponding author had full access to all the data in the study and had final responsibility for the decision to submit for publication.

## RESULTS

Between November 2016 and July 2019, 2376 individual WLH were enrolled in the study including 1217 enrolled prior to randomization (Figure 1). Characteristics of participants did not differ by enrollment period (supplemental Table 1). Overall, 1900 participants had at least one follow-up visit or were enrolled after randomization and were included in this analysis of the effect of CQI, including, 1103 (58.0%) from the control health zones and 797 (42.0%) from health zones in the intervention group (Table 1). Overall, compared with the control group, clinics in the intervention group tended to be more peri-urban (51·8% vs 29·4%, p <.0001), to receive technical support from a PEPFAR implementing partner (70·1% vs 57·8%, p <.0001), unlikely to be managed by the government (22·2% vs 24·2%, p <.0001) (Table 1). At individual level, participants in the control group had a slightly higher median years of education (12 vs 11 years, p= 0·0001), were more likely to be married or cohabitating (72·3% vs 64·0%, p <.0001), primigravida (12.0% vs 8.8%, p= 0·0258) (Table 1).

**Figure 1.**
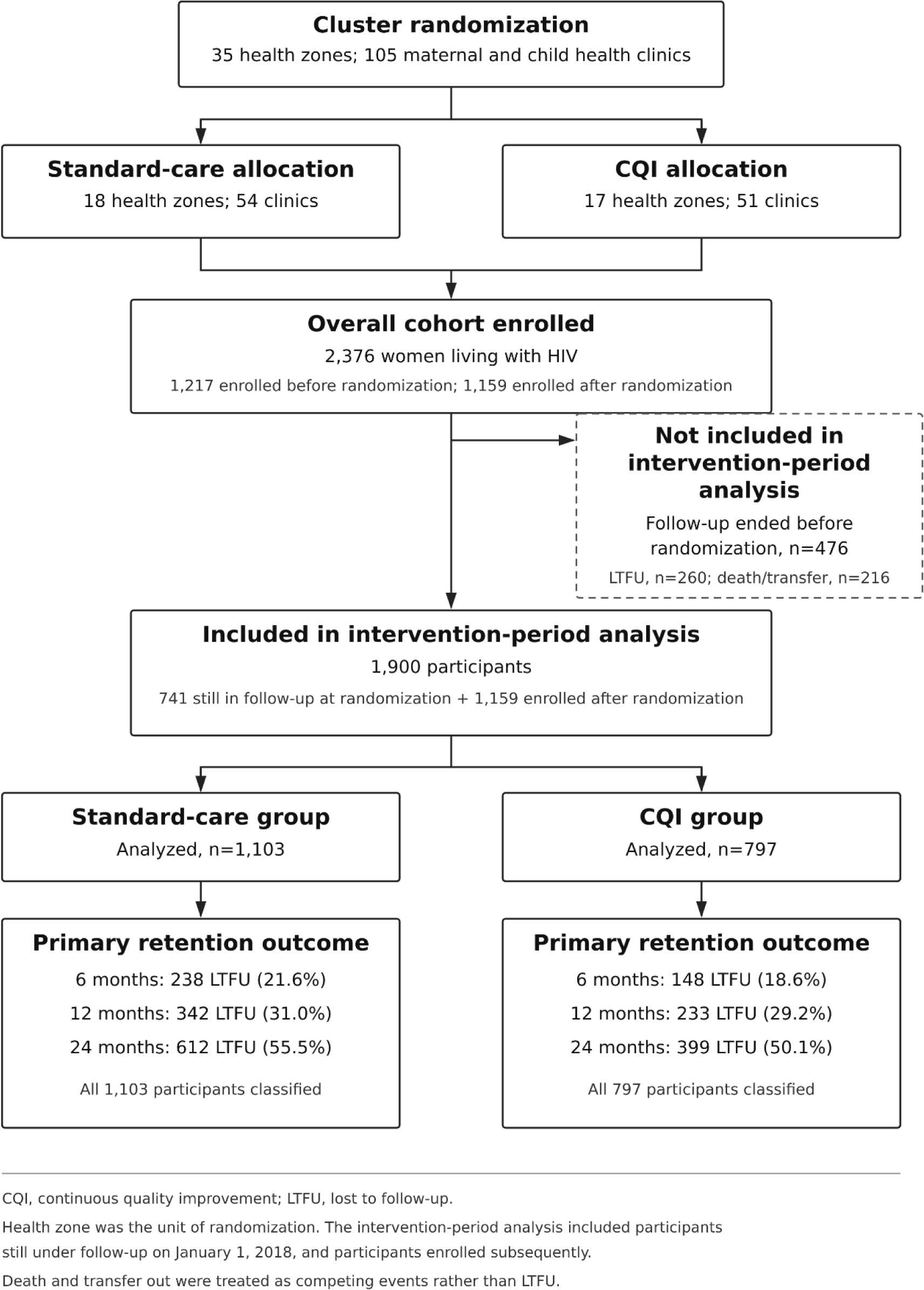
Flow of clusters and participants through the trial

**Table 1.** Individual and clinical characteristics of 1900 participants included in the intervention-period analysis by randomization group.

|  | Intervention Phase |  |  | P-value<sup>§</sup> |
| --- | --- | --- | --- | --- |
|  | Overall | Control | Intervention |  |
|  | Number (%) | Number (%) | Number (%) |  |
| **Age in years: median (IQR)** | 31 (27, 36) | 31 (27, 36) | 32 (27, 36) | 0.2107 |
| ≤ 24 | 289 (15.23) | 177 (16.08) | 112 (14.05) | 0.4615 |
| 25 - 34 | 1007 (53.06) | 581 (52.77) | 426 (53.45) |  |
| 35 + | 602 (31.72) | 343 (31.15) | 259 (32.50) |  |
| Missing | 2 | 2 | 0 |  |
| **Maternal education** |  |  |  |  |
| Years: median (IQR) | 11 (9, 12) | 12 (9, 12) | 11 (9, 12) | <0.0001 |
| primary | 241 (12.72) | 132 (12.01) | 109 (13.69) | 0.002 |
| secondary | 1361 (71.82) | 770 (70.06) | 591 (74.25) |  |
| tertiary | 293 (15.46) | 197 (17.93) | 96 (12.06) |  |
| Missing | 5 | 4 | 1 |  |
| **marital status** |  |  |  |  |
| Married/ Live-in boyfriend | 1316 (69.37) | 806 (73.27) | 510 (63.99) | <0.0001 |
| Never married/separated/divorced | 581 (30.63) | 294 (26.73) | 287 (36.01) |  |
| Missing | 3 |  |  |  |
| **Any occupation** |  |  |  |  |
| No | 1180 (62.11) | 652 (59.11) | 528 (66.25) | 0.0016 |
| Yes | 720 (37.89) | 451 (40.89) | 269 (33.75) |  |
| **Disclosure of HIV status** |  |  |  |  |
| Non-disclosed | 876 (46.15) | 491(44.60) | 385 (48.31) | 0.1095 |
| disclosed | 1022 (53.85) | 610 (55.40) | 412 (51.69) |  |
| Missing | 2 |  |  |  |
| **primigravida** |  |  |  |  |
| Yes | 202 (10.64) | 132 (11.98) | 70 (8.78) | 0.0258 |
| No | 1697 (89.36) | 970 (88.02) | 727 (91.22) |  |
| Missing |  |  |  |  |
| **Transportation mean** |  |  |  |  |
| Walking | 700 (36.86) | 396 (35.93) | 304 (38.14) | 0.3249 |
| Taxi or other | 1199 (63.14) | 706 (64.07) | 493 (61.86) |  |
| **Time of Enrollment** |  |  |  |  |
| during pregnancy | 737 (38.79) | 421 (38.17) | 316 (39.65) | 0.5135 |
| Delivery or postpartum | 1163 (61.21) | 682 (61.83) | 481 (60.35) |  |
| **Duration of ART at enrollment** |  |  |  |  |
| < 12 months | 885 (46.73) | 500 (45.58) | 385 (48.31) | 0.0735 |
| 13-24 months | 184 (9.71) | 97 (8.84) | 87 (10.92) |  |
| >= 24 months | 825 (43.56) | 500 (45.58) | 325 (40.78) |  |
| Missing | 6 |  |  |  |
| **Time of diagnosis** |  |  |  |  |
| During this pregnancy | 800 (42·13) | 448 (40·65) | 352 (44·17) | 0·1261 |
| prior to this pregnancy | 1099 (57·87) | 654 (59·35) | 445 (55·83) |  |
| Missing | 1 |  |  |  |
| **Management Type** |  |  |  |  |
| Government/public | 444 (23·37) | 267 (24·21) | 177 (22·21) | <·0001 |
| Faith-based | 636 (33·47) | 289 (26·20) | 347 (43·54) |  |
| Private | 820 (43·16) | 547 (49·59) | 273 (34·25) |  |
| **PEPFAR Support** |  |  |  |  |
| No | 704 (37·05) | 466 (42·25) | 238 (29·86) | <·0001 |
| Yes | 1196 (62·95) | 637 (57·75) | 559 (70·14) |  |
| **Location** |  |  |  |  |
| Urban | 1162 (61·22) | 779 (70·63) | 383 (48·18) | <·0001 |
| peri-urban | 736 (38·78) | 324 (29·37) | 412 (51·82) |  |
| Missing | 2 |  |  |  |

### Lost to follow-up at 6-, 12-, and 24 months post randomization/enrollment

Of the 1900 WLH in care at randomization or enrolled after, 386 (20·3%), 575 (30·3%), and 1011 (53·2%), respectively, were LTFU at 6, 12, and 24 months after randomization/enrollment. Across all three time points, participants in the intervention group were consistently less likely to be lost to follow-up: 18·6% vs 21·6% (aRD −5.2%; 95%CI: −9.6%, −0.8%), 29·2% vs 31·0% (aRD −3.9%; 95%CI: −10.2%, 2.4%), and 50·1% vs 55·5%; (aRD −7.1%; 95%CI: −15.2%, 1.0%), respectively (Table 2).

**Table 2.** Effect of CQI on lost-to-follow-up at 6, 12, and 24 months after randomization.

|  |  |  |  |  | Risk difference (95%CI)<sup>1</sup> |  |
| --- | --- | --- | --- | --- | --- | --- |
|  | Total | LTFU | Dead | Transfer | Crude | Adjusted<sup>1</sup> |
| **6 months** |  |  |  |  |  |  |
| Control | 1103 | 238 (21.6) | 3 (0.27) | 68 (6.17) |  |  |
| Intervention | 797 | 148 (18.57) | 5 (0.63) | 39 (4.89) | -3.0%(-9.9%, 3.9%) | -5.2% (-9.6%, -0.8%) |
| **12 months** |  |  |  |  |  |  |
| Control | 1103 | 342 (31.01) | 5 (0.45) | 80 (7.25) |  |  |
| Intervention | 797 | 233 (29.23) | 6 (0.75) | 42 (5.27) | -1.8% (-10.6%, 7.0%) | -3.9% (-10.2%, 2.4%) |
| **24 months** |  |  |  |  |  |  |
| Control | 1103 | 612 (55.49) | 7 (0.63) | 94 (8.52) |  |  |
| Intervention | 797 | 399 (50.06) | 7 (0.88) | 46 (5.77) | -5.4% (-15.8%, 4.9%) | -7.1% (-15.2%, 1.0%) |

<sup>1</sup> Adjusted for marital status, educational level, occupation, primigravida, management of the structure, urban/peri-urban location, implementation supporting partner. Estimates are obtained using binomial regression model with an identity link.

When analysis was stratified by urban vs peri-urban location, the intervention has virtually no effect on LTFU in peri-urban area: aRD −3.8%; 95%CI: −10.5%, 2.9%) at 6 months, 2.4%; 95%CI: −7.6%, 12.3%) at 12 months, and 2.7%; 95%CI: −6.8%, 12.2%) at 24 months, respectively. In urban areas, the effect of the intervention was stronger: aRD −7.2%; 95%CI: −12.6%, −1.8%) at 6 months, −7.2%; 95%CI: −14.1%, −0.3%) at 12 months, and −10.2%; 95%CI: −19.3%, −0.8%) at 24 months respectively. Similar difference was observed when stratification by receipt of direct PEPFAR support with virtually no effect of the intervention in non-PEPFAR facilities and a strong effect in PEPAR supported facilities (supplemental Table 2). Although point estimates suggested a potentially greater intervention effect in urban and PEPFAR-supported facilities, the interaction tests were not statistically significant; these subgroup findings should therefore be interpreted cautiously.

In the secondary analyses, Gray’s test showed no evidence of a difference between groups during the preintervention period (p=0.7882), whereas the cumulative incidence functions differed during the postintervention period (p=0.0002). The curves began to separate early and generally diverged further over time (Figure 2), suggesting a sustained intervention benefit.

**Figure 2a.**
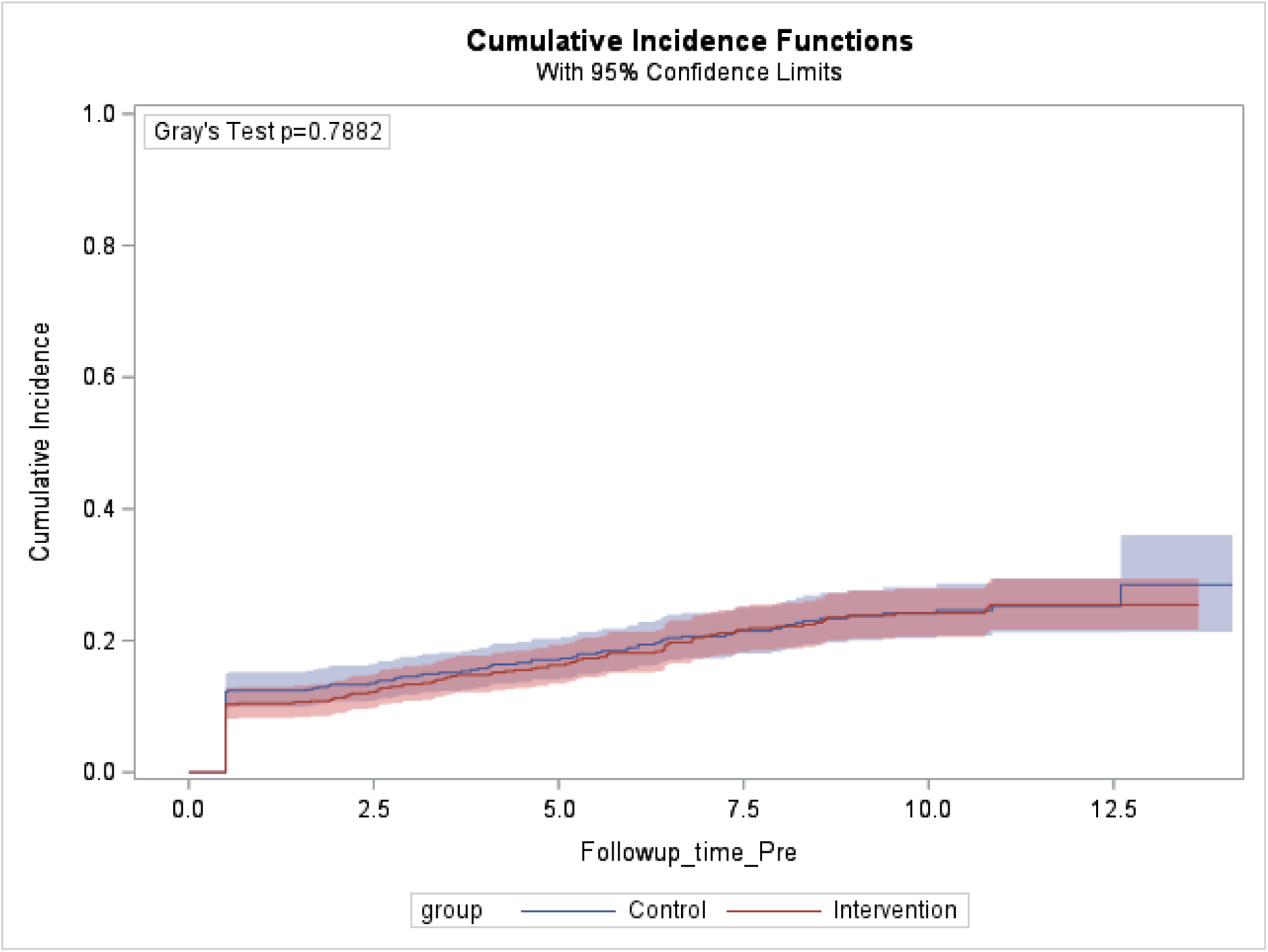
Cumulative lost-to-follow-up in the pre-intervention period by group.

**Figure 2b.**
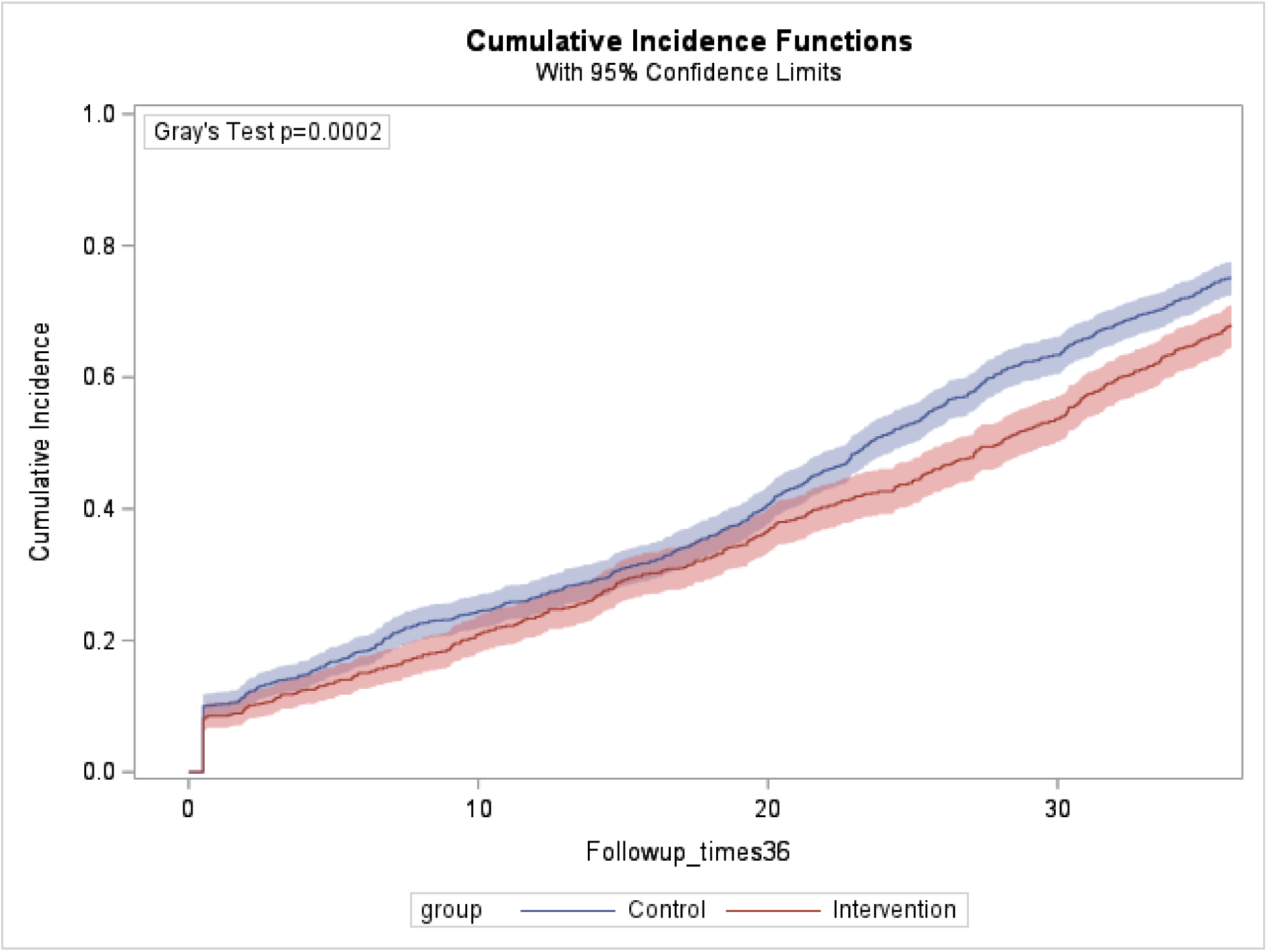
Cumulative lost-to-follow-up in the post-intervention period by intervention group.

The facility-level variance at 6 months was estimated at 0.2998, yielding a facility ICC of 0.0835, 95%CI: 0.0245, 0.1358 (supplemental Table 3).

### Viral Suppression at 0, 6-, 12-, and 24 months post randomization/enrollment

Of the 1900 eligible WLH, 1145 (60·3%) had a viral load result within 6 months prior or 3 months after randomization/enrollment. Of the 1399, 1192, 735 in care at 6-, 12- and 24 months post randomization/enrollment, 534 (38·2%), 601 (50·4%), and 540 (73·5%), respectively, had a viral load result. Availability of viral load results did not differ substantially by intervention groups (Table 3). Among those with available viral-load result, 782 (68·3%) at randomization/enrollment, 394 (73·8%) at 6 months, 470 (78·2%) at 12 months, and 408 (75·6%) at 24 months were virologically suppressed. The proportions of participants with virological suppression did not differ by randomization groups at any time point (Table 3).

**Table 3.**
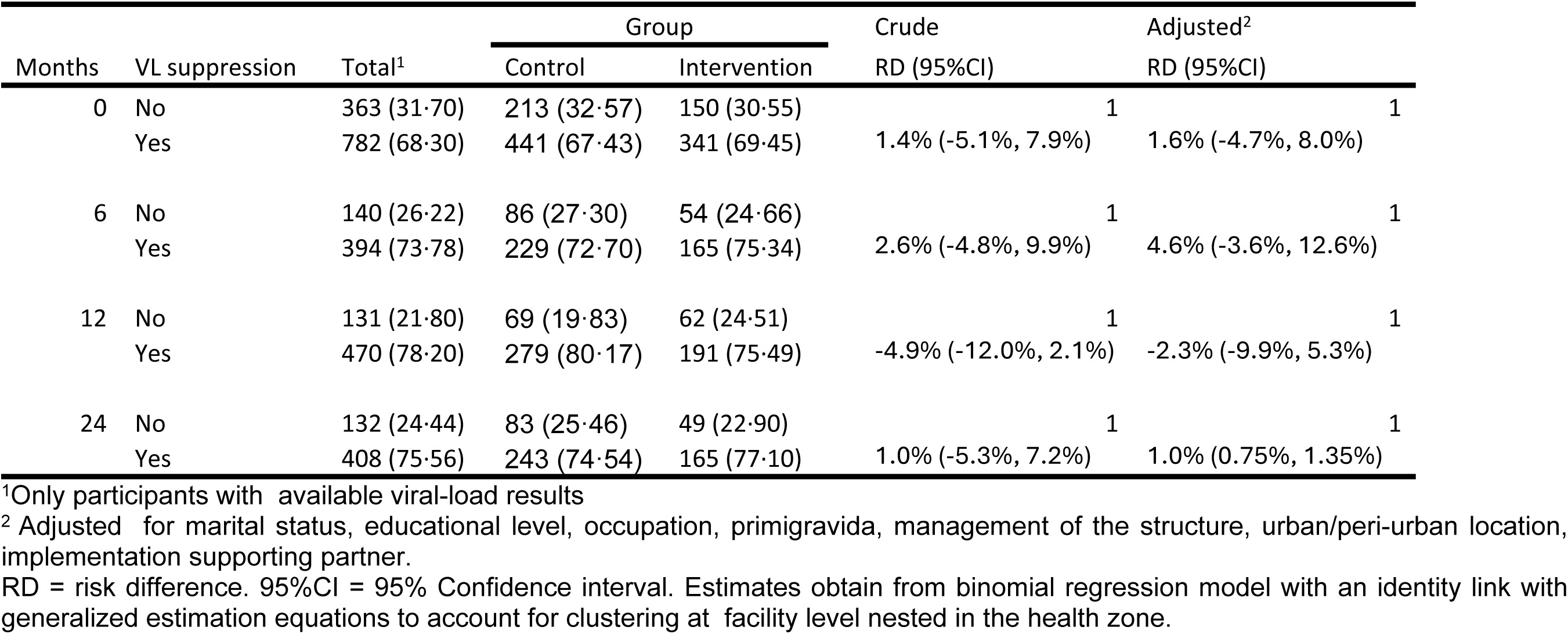
Effect of CQI on viral suppression at 6, 12, and 24 months after randomization.

### Early infant diagnosis and vertical transmission

Of the 1900 women in care or enrolled after randomization, 1549 had a live birth and at least one follow-up visit during the intervention period. Among their children, over 88% of those seen at 6 weeks have a blood sample collected for early infants’ diagnosis and a similar proportion at 12 months for serology, with no difference by randomization group (Table 4). However, results of these tests were not immediately available. At the closure of the data for this analysis, only 125 (8·1%) had a PCR DNA test result available at 6 weeks, 507 (32·7%), 529 (34·2%), 492 (31·8%), 442 (28·5%) at 6, 12, 18, and 24 months, respectively. Infant diagnosis test results were more frequently available in the control group (Table 4).

**Table 4.**
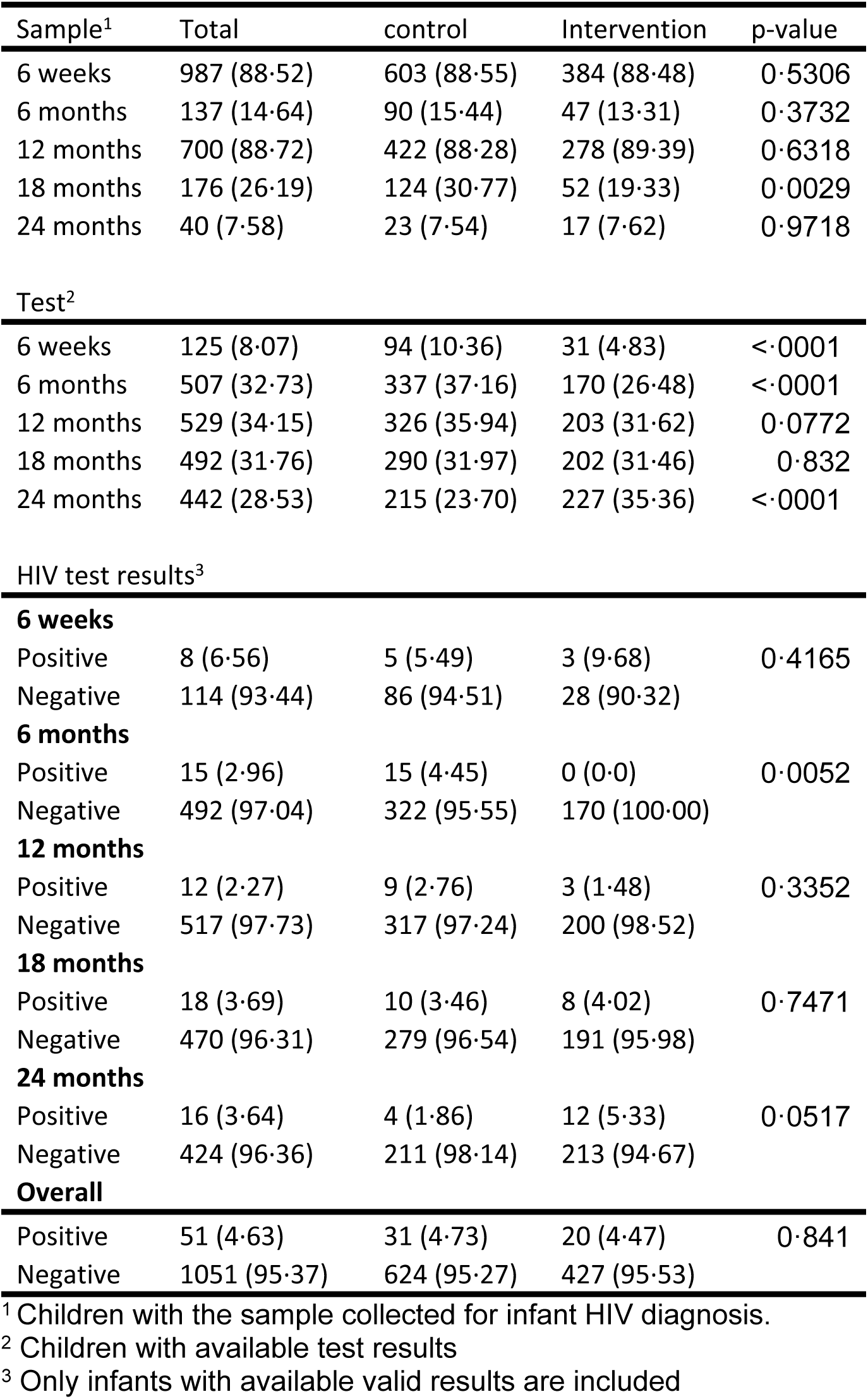
Infant HIV diagnosis testing and results at 6 weeks, 6-, 12-, 18-, 24 months after delivery.

### CQI implementation across facilities

Data from the one-year postintervention implementation survey was available for 50/51 facilities (one survey response was lost during the translation and transcription process). All facilities reported having a CQI team (Table 5). Six (12%) facilities reported holding no CQI meeting, while 36 (72%) reported monthly meetings or more as per protocol. At least one CQI meeting report was available in 26 (52%) facilities and all facilities reported regular supervisory visits. Among the 50 facilities, 43 (86%) reported at least one challenge in implementing their CQI initiatives. The most frequently reported challenges were stock-outs or delayed availability of essential supplies and difficulties tracing participants because of incorrect addresses, unreachable telephone numbers, distance, or transportation barriers. Facilities also reported limited staff and CQI-team engagement, inadequate training or unclear responsibilities, insufficient financial and logistical support, challenges engaging male partners, and participant-level barriers related to HIV-status acceptance, disclosure, stigma, and religious beliefs (Table 5).

**Table 5.**
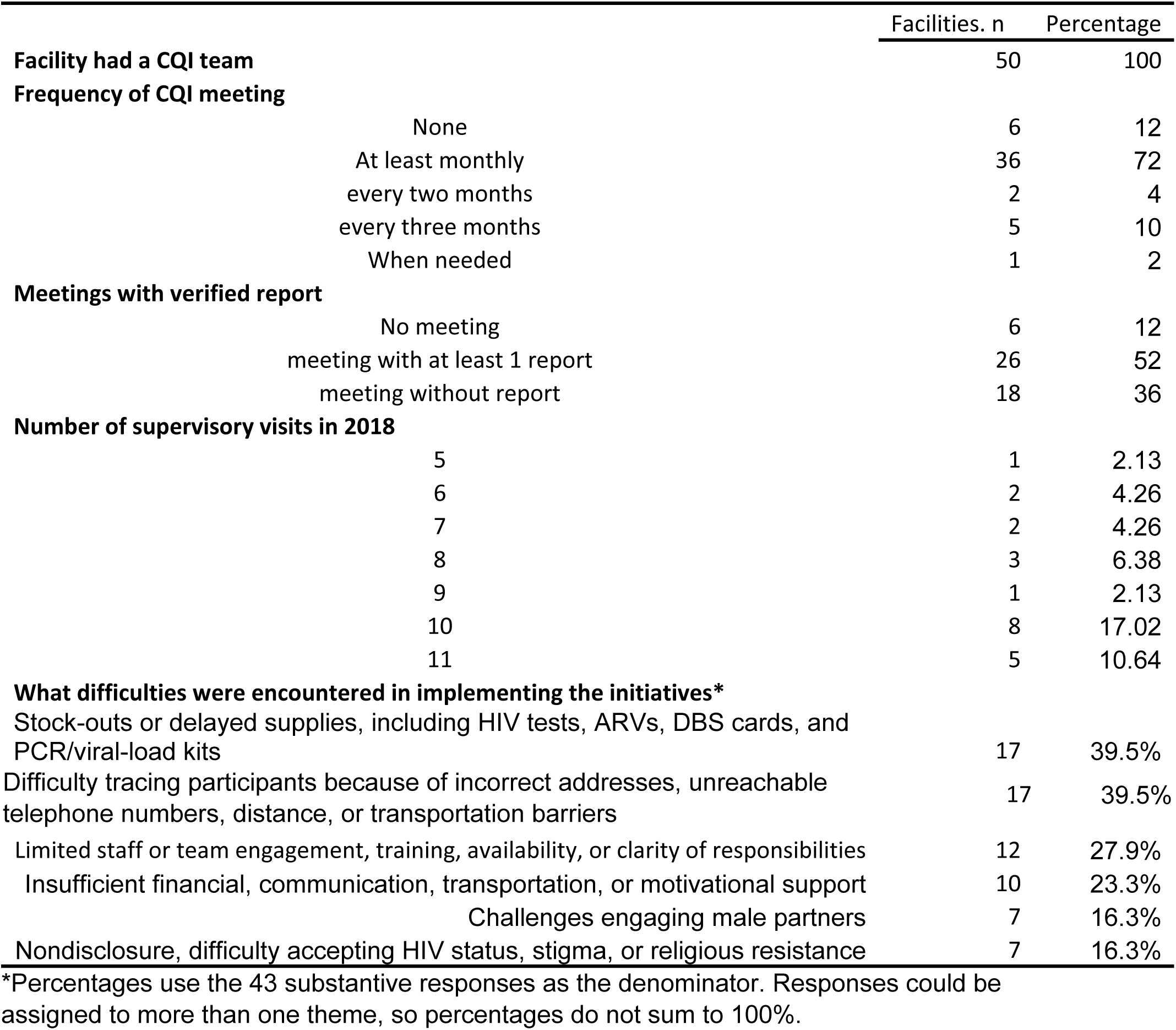
Results of the mid-term evaluation of CQI implementation.

## DISCUSSION

In this pragmatic cluster randomized controlled study, we aimed to evaluate the effectiveness of data-driven continuous quality improvement (CQI) implementation on LTFU, virological outcomes, and early infant diagnosis among pregnant and breastfeeding WLH and their HIV-exposed infants receiving ART in MCH clinics in Kinshasa, DRC. We found that CQI modestly reduced loss-to-care by approximately −5, −4, and −7 percentage points at 6, 12, and 24 months, respectively. The reduction was statistically significant at 6 months. For patients retained in care, viral load results were only available for 60%, 38%, 50%, 74% of participants at enrollment/randomization, 6-, 12-, and 24 months. Among those with available viral load, the proportion with suppressed viral loads ranged from 68% at baseline, to 74%,78%, and 76% at 6-, 12-, and 24 months, respectively, and did not differ by intervention groups. Similarly, although close to 90% of infants had documented blood sample collection for HIV testing at different evaluation time points, results were available only for 8% of infants at 6 weeks postpartum, 33%, 34%, 32% and 29% at 6-, 12-, 18-, and 24 months, respectively, and more likely in the control group.

To our knowledge, the only trial to evaluate the effectiveness of CQI interventions on LTFU among pregnant and breastfeeding women is the trial from Nigeria. At 6 months postpartum the increase in retention in the intervention group was about 3%. Although not statistically significant, this effect size is similar to the effect size from our own study. Further, though the trial in Nigeria reported rates of early infant testing at 4–6 weeks were higher in intervention sites (48·8% vs. 25·3%, adjusted relative risk: 1·76; 95% CI: 1·27 to 2·42) [22], because of the challenges in obtaining results from the National Reference laboratory, this outcome could not be evaluated in our study despite a high rate of sample collection. In a second study that used a stepped-wedge cluster-randomized controlled implementation trial to evaluate the effectiveness of continuous quality improvement (CQI) on viral load (VL) monitoring and repeat HIV testing in rural South Africa, CQI implementation was found to significantly increase VL monitoring (relative risk [RR] 1·38, 95% CI 1·21 to 1·57, p < 0·001) but did not improve repeat HIV testing [23]. In our study, again limited by the availability of viral load results, we did not find any differences in viral load outcomes by group.

System approaches to improve health service outcomes such as CQI are based on the premise that the poor outcomes are the result of poor health service delivery that could be addressed if timely identified and the attention of the appropriate provider is focused on it. In our study, CQI produced a modest improvement in retention, an outcome that clinic teams could directly influence through activities such as monitoring missed visits and contacting participants who were late for care. By contrast, viral-load monitoring and infant HIV diagnosis depended on processes extending beyond participating clinics and health-zone teams. Although specimen collection was relatively high, testing and the timely return of results relied on the national reference laboratory, which was outside the intervention’s operational reach. These findings suggest that local quality-improvement interventions may strengthen processes within frontline teams’ control but cannot alone overcome failures elsewhere in an interconnected care system. Future interventions should map the full service-delivery cascade and align their operational reach with all health-system levels responsible for the intended outcomes.

Retention in care, particularly in the postpartum period, is a major obstacle to achieving the goals of ending AIDS by 2030 for children 0-14 years of age. Our study provides randomized evidence that CQI interventions can be used to improve retention in care for up to 24 months (i.e at the end of the breastfeeding period) and support the WHO’s call for the development and promotion of quality improvement methods to strengthen regional and district-level health systems to improve the quality and reliability of HIV prevention, care and treatment services for women and children.

This study had several limitations. First, because randomization was at the health zone it was difficult to stratify. One consequence is that the groups differed in participant numbers and several baseline characteristics. But adjustment for these variables did not change the effect size of CQI. Second, implementation of CQI interventions was the responsibility of the site. The only support provided by the study was logistics for data collection, production of indicators, and for the organization of quarterly meetings. As such, variations in the level of intensity of engagement by sites were to be expected. However, given the limited number of participants at many sites, it was not possible to analyze the effect of CQI by sites. Third, despite efforts to avoid contamination, the study was implemented in collaboration with the provincial coordination of the National AIDS Program which has oversight on all HIV programs in the province, it is not impossible that clinics in the control health zone learned about the intervention and adopted some aspects of it. The overall improvement in the availability of viral load over the follow-up time is indirect evidence of this potential contamination. Fourth, the selection of virological suppression and infant HIV testing as study’s end points when the main issue related to these outcomes was at the level of national reference laboratory and the study intervention was not designed to reach the laboratory. Design of CQI intervention should carefully examine the cascade and select end points that are directly addressed by the interventions. Finally, only 52% of facilities had documented CQI meeting reports, indicating variable implementation fidelity.

Strengths of this study include its pragmatic design, its implementation in a resource-limited setting and high- and low-volume clinics, suggesting that the results may be generalizable to similar contexts. The pragmatic nature of the study and the low-intensity made sustainability easy. Although we did not collect systematic data on this, towards the end of the study, many facilities were well versed in exploiting their own data to identify points of intervention.

In conclusion, data-driven CQI implementation in MCH clinics in Kinshasa produced a modest improvement in retention, with the clearest evidence at 6 months and favorable but imprecise estimates through 24 months. Limited availability of maternal viral-load and infant HIV-test results precluded definitive conclusions about virological suppression and vertical transmission. Local CQI may strengthen processes within frontline teams’ control, but achieving broader outcomes will require interventions whose operational reach extends across all health-system levels responsible for those outcomes.

## Contributors

MY designed the study. MY and PB obtained funding, MY, NLRR, BK, MT, FM, FL, NZ, and PB implemented the study and collected data. MY analyzed the data and wrote the first draft of the report. All authors contributed to the final report.

## CONFLICTS OF INTEREST

The authors declare no competing interests

## ACKNOWLEDGEMENTS

We are grateful for the participation and time of the mothers and infants in the study, the time and efforts of the personnel of the participating clinics and health zone bureaus. We acknowledge the contribution of the following site investigators of the CQI-PMTCT study team: Godelive Aitikalema, Ali Alisho, Elysée Bayayana, Alain Bilwaya, Fabrice Bumwana, Pierre Dianzenza, Jean Claude Dinanga, Grace Kabangu, André Kapuku, Georges Kihuma, Willy Lukumu, Fidèle Lumande, Zouzou Masevo, Fanny Matadi, Rachel Mushiya, Marie Therèse Mwela, Willy Ndala, Justin Ngelengele, Patrick Ngimbi, José Nlandu, Huguette Mbali, Pearl Tenatena and Marie Tshibuabua, Emile Okitolonda, Sandra Osuku, Frieda Behets, and the support of Einstein College of Medicine’s, the Ohio State University’s, and Kinshasa School of Public Health’s administrative teams.

## Data availability statement

The individual-level data underlying this study cannot be made publicly available because they contain sensitive information concerning HIV status, pregnancy, and clinical care, and public data sharing was not included in the participants’ informed-consent process. A deidentified analytic dataset, accompanying data dictionary, and statistical analysis code are available to qualified researchers upon reasonable request and subject to approval by the Albert Einstein Institutional Review Board (IRB). Requests should be submitted to and should include a brief research proposal, analysis plan, institutional affiliation, and documentation of applicable ethical approval. Approved researchers will be required to sign a data-use agreement protecting participant confidentiality and limiting use of the data to the approved research purpose. Statistical analysis code available upon request

## Funding

This research was supported by the President’s Emergency Plan for AIDS Relief (PEPFAR) and the Eunice Kennedy Shriver National Institute of Child Health and Human Development (NICHD R01HD087993). FL, NZ, PB, and MY are partially supported by the National Institutes of Health (NIAID U01AI096299, NIHCD R01HD105526). The sponsors of the study had no role in study design, data collection, data analysis, data interpretation, writing of the report, or the decision to submit the paper for publication.

## ICC calculation

To quantify the degree of clustering of LTFU within health facilities, we fitted a logistic generalized linear mixed-effects model with a random intercept for facility nested within health zone. The facility-level intraclass correlation coefficient (ICC) was calculated on the latent logistic scale as 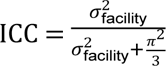 where 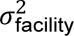 represents the estimated variance of the facility-level random intercept and *π*^2^/3 is the assumed residual variance of the logistic distribution. Thus, the ICC represents the proportion of the total latent-scale variance attributable to differences between facilities. The 95% confidence interval (CI) for the ICC was obtained by first calculating a Wald 95% CI for the facility-level variance component and then transforming the lower and upper variance limits using the same ICC formula, with the lower variance bound constrained to zero.

---

**Supplemental Table 1.** Individual and clinical characteristics by enrollment before or after randomization.

|  | Overall | Pre-randomization | After Randomization | P-value<sup>6</sup> |
| --- | --- | --- | --- | --- |
|  | N (%) | N (%) | N (%) |  |
| **Age in years: median (IQR)** | 31 (27, 36) | 32 (27, 36) | 31 (27, 36) | 0.6853 |
| <= 24 | 379 (156.1) | 190 (15.6) | 189 (16.4) | 0.8571 |
| 25 - 34 | 1250 (52.7) | 641 (52.7) | 609 (52.7) |  |
| 35 + | 743 (31.3) | 386 (31.7) | 357 (30.9) |  |
| Missing | 3 | 1 | 2 |  |
| **marital status** |  |  |  |  |
| Married/ Live-in boyfriend | 1623 (68.4) | 375 (30.8) | 373 (32.3) | 0.4464 |
| Never married/separated/divorced | 748 (31.6) | 842 (69.2) | 781 (67.7) |  |
| Missing | 3 |  |  |  |
| **Maternal education** |  |  |  |  |
| Years: median (IQR) | 11 (9, 1) | 11 (8, 1) | 11 (9, 1) | 0.0122 |
| primary | 309 (13.1) | 177 (14.6) | 132 (11.5) | 0.0015 |
| secondary | 1700 (71.8) | 851 (70.0) | 849 (73.7) |  |
| tertiary | 359 (15.2) | 188 (15.5) | 171 (14.8) |  |
| Missing | 6 | 1 | 5 |  |
| **Anny Occupation** |  |  |  |  |
| No | 1494 (62.9) | 762 (62.6) | 732 (63.3) | 0.7502 |
| Yes | 880 (37.1) | 455 (37.4) | 425 (36.7) |  |
| Missing |  |  |  |  |
| **Disclosure of HIV status** |  |  |  |  |
| nondisclosed | 1137 (48.1) | 594 (49.1) | 543 (47.0) | 0.3336 |
| disclosed | 1228 (51.9) | 616 (50.9) | 612 (53.0) |  |
| Missing | 9 | 7 | 2 |  |
| **primigravida** |  |  |  |  |
| Yes | 250 (10.5) | 112 (9.2) | 138 (11.9) | 0.0323 |
| No | 2123 (89.5) | 1104 (90.8) | 1019 (88.1) |  |
| Missing | 1 | 1 |  |  |
| **Transportation means** |  |  |  |  |
| Walking | 1483 (62.5) | 456 (37.5) | 722 (62.5) | 0.9616 |
| Taxi or other | 890 (37.5) | 761 (62.5) | 434 (37.5) |  |
| Missing | 1 |  |  |  |
| **Time of Enrollment** |  |  |  |  |
| during pregnancy | 1390 (58.5) | 591 (48.6) | 764 (66.0) | <.0001 |
| Delivery or postpartum | 984 (41.5) | 626 (51.4) | 393 (34.0) |  |
| Missing |  |  |  |  |
| Duration of ART at enrollment |  |  |  |  |
| < 12 months | 1130 (47.7) | 544 (44.7) | 586 (50.9) | 0.0004 |
| 13-24 mths | 235 (9.9) | 145 (11.9) | 90 (7.8) |  |
| >= 24 mths | 1003 (42.4) | 528 (43.4) | 475 (41.3) |  |
| Missing | 6 |  | 6 |  |
| Time of diagnosis |  |  |  |  |
| During this pregnancy | 1041 (43.9) | 529 (43.5) | 512 (44.3) | 0.6518 |
| prior to this pregnancy | 1332 (56.1) | 688 (56.5) | 644 (55.7) |  |
| Missing | 1 |  | 1 |  |
| Management Type |  |  |  |  |
| Government/public | 571 (24.0) | 299 (24.6) | 272 (23.5) | 0.6317 |
| Faith based | 769 (32.4) | 384 (31.5) | 385 (33.3) |  |
| Private | 1034 (43.6) | 554 (43.9) | 500 (43.2) |  |
| Missing |  |  |  |  |
| PEPFAR Support |  |  |  |  |
| No | 908 (38.3) | 456 (37.5) | 452 (39.1) | 0.443 |
| Yes | 1466 (61.7) | 761 (62.5) | 705 (60.9) |  |
| Missing |  |  |  |  |
| Location |  |  |  |  |
| Urban | 1482 (62.5) | 778 (64.0) | 704 (60.8) | 0.107 |
| peri-urban | 890 (37.5) | 437 (36.0) | 453 (39.2) |  |
| Missing | 2 | 2 |  |  |

<sup>δ</sup>Comparing intervention group with control group: Wilcoxon rank-sum test for continuous variable and Pearson Chi Square for categorical variable

**Supplemental table 2.** Effect of CQI on viral suppression at 6, 12, and 24 months after randomization stratified by facility location or receipt of technical support from a PEPFAR implementing partner.

|  | peri-urban |  | Urban |  |
| --- | --- | --- | --- | --- |
|  | Crude | adjusted | Crude | adjusted |
| **6 months** |  |  |  |  |
| Control |  |  |  |  |
| Intervention | 0.9% (-10.4%, 12.2%) | -3.8% (-10.5%, 2.9%) | -5.1% (-11.4%, 1.2%) | -7.2% (-12.6%, -1.8%) |
| **12 months** |  |  |  |  |
| Control |  |  |  |  |
| Intervention | 4.9% (-10.1%, 19.9%) | 2.4% (-7.6%, 12.3%) | -5.4% (-12.4%, 1.5%) | -7.2% (-14.1%, -0.3%) |
| **24 months** |  |  |  |  |
| Control |  |  |  |  |
| Intervention | -1.5% (-19.8%, 16.8%) | 2.7% (-6.8%, 12.2%) | -8.2% (-17.3%, 0.9%) | -10.2% (-19.3%, -0.8%) |
| **6 months** | Non PEPFAR |  | PEPFAR |  |
| Control |  |  |  |  |
| Intervention | -0.4% (-8.9%, 8.1%) | -0.9% (-7.4%, 5.7%) | -4.8% (-14.3, 4.7%) | - |
| **12 months** |  |  |  |  |
| Control |  |  |  |  |
| Intervention | 0.9% (-9.4%, 11.1%) | 0.1% (-7.6%, 7.8%) | -3.5% (-15.6%, 8.5%) | -7.7% (-15.8%, 0.4%) |
| **24 months** |  |  |  |  |
| Control |  |  |  |  |
| Intervention | -0.2% (-10.4%, 10.0%) | -0.2% (-9.2%, 8.8%) | -9.4% (-23.2%, 4.5%) | -11.4% (-21.9%, -0.8%) |

<sup>1</sup>Adjusted for marital status, educational level, occupation, primigravida, management of the structure, urban/peri-urban location, implementation supporting partner. RD = Risk difference, 95%CI = 95% confidence interval.

**Supplemental Table 3.**
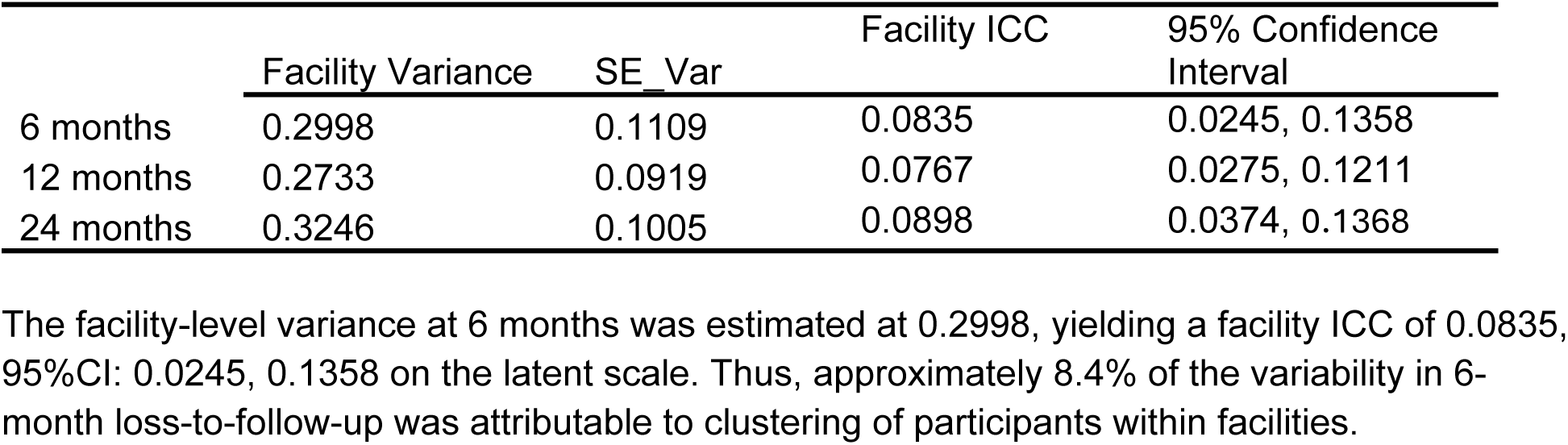
Intracluster correlation coefficients for Facility at each timepoint for LTFU.

|  | Facility Variance | SE Var | Facility ICC | 95% Confidence Interval |
| --- | --- | --- | --- | --- |
| 6 months | 0.2998 | 0.1109 | 0.0835 | 0.0245, 0.1358 |
| 12 months | 0.2733 | 0.0919 | 0.0767 | 0.0275, 0.1211 |
| 24 months | 0.3246 | 0.1005 | 0.0898 | 0.0374, 0.1368 |

The facility-level variance at 6 months was estimated at 0.2998, yielding a facility ICC of 0.0835, 95%CI: 0.0245, 0.1358 on the latent scale. Thus, approximately 8.4% of the variability in 6-month loss-to-follow-up was attributable to clustering of participants within facilities.

## Notes

### Competing Interest Statement

The authors have declared no competing interest.

### Clinical Trial

NCT03048669

### Clinical Protocols

https://pubmed.ncbi.nlm.nih.gov/28446232/

